# A polygenic risk score for rheumatoid arthritis sheds light on the Prevotella association

**DOI:** 10.1101/2019.12.09.19014183

**Authors:** Philippa M Wells, Adewale S Adebayo, Maxim B Freidin, Axel Finckh, Till Strowig, Till Robin Lesker, Deshire Alpizar-Rodriguez, Benoit Gilbert, Bruce Kirkham, Andrew P Cope, Claire J Steves, Frances M K Williams

**Affiliations:** Department of Twin Research and Genetic Epidemiology, King’s College London, London, UK; Division of Rheumatology, Geneva University Hospital, Geneva, Switzerland; Department of Microbial Immune Regulation, Helmholtz Centre for Infection Research, Braunschweig, Germany; Hannover Medical School, Hannover, Germany; Department of Rheumatology, Guy’s and St Thomas’ NHS Trust, London, UK; Centre for Rheumatic Diseases, King’s College London, London, UK; Centre for Inflammation Biology and Cancer Immunology, King’s College London, London, UK; Department of Ageing and Health, St Thomas’ Hospital, London, UK

**Keywords:** *Prevotella*, rheumatoid arthritis, host genotype, microbiome, inflammation

## Abstract

**Background:** Rheumatoid arthritis (RA) is a chronic inflammatory autoimmune disease associated with reduced life expectancy. It is heritable and an extensive repertoire of genetic variants have been identified. The gut microbiota may represent an environmental risk factor for RA. Indeed, *Prevotella copri* is a candidate keystone species, but whether it lies on the causal pathway for disease or is simply a bystander reflecting host-genetic predisposition to RA, remains to be determined. The study of disease-microbiota associations may be confounded by the presence of the disease of interest or by its treatment. To circumvent this issue, we assessed whether known RA risk alleles were associated with the gut microbiota, in a large who do not have RA.

**Methods:** Blood and stool acquired from volunteers from TwinsUK were used for genotyping and assessment of the gut microbiota, respectively. A weighted polygenic risk score (PRS) for RA was calculated in 1650 unaffected twins from the TwinsUK cohort, based on 233 published GWAS-identified published RA associated single nucleotide polymorphisms (SNPs). Amplicon sequence variants (ASVs) were generated from 16S rRNA sequencing of stool samples and assessed for association with RA PRS. Confirmation of findings was performed using an independent sample comprised of first-degree relatives of RA patients from the SCREEN-RA cohort (n=133).

**Findings:** We found that *Prevotella spp* was positively associated with RA PRS in TwinsUK participants (p.adj<1e-7). This finding was validated in SCREEN-RA participants carrying the shared epitope risk alleles (p.adj=1.12e-3). An association of *Prevotella spp*. with pre-clinical RA phases was also demonstrated (p.adj=0.021).

**Interpretation:** *Prevotella* in the gut microbiota is associated with RA genotype in the absence of RA, as well as in subjects at high risk of developing RA. This work suggests that host genotype is associated with microbiota profile prior to disease onset.

**Research in context:** *Evidence before this study?:* *Prevotella copri* is increased within the gut microbiota in rheumatoid arthritis (RA) patients, predominantly those with early disease, before treatment is initiated. *Prevotellaceae* (family) has been demonstrated to be increased in pre-clinical RA cases compared to controls. *Prevotella copri* is posited to be an inflammatory driver, contributing to RA pathology by promoting a pro-inflammatory cytokine milieu.

*What does this study add?:* This study is the first to demonstrate that, in a large human cohort, carrying genetic risk factors for RA is associated with a higher abundance of *Prevotella spp*. in the absence of RA.

*How might this impact on clinical practice or future developments:* This work suggests that any potential causal role of *Prevotella spp*. occurs very early in disease development. Strategies to intervene should target early or asymptomatic at-risk groups

## Introduction

Rheumatoid arthritis (RA) is a debilitating chronic autoimmune condition, associated with reduced life expectancy. There is a substantial genetic component to RA aetiology, with heritability estimated at 65%.^1^ Known environmental risk factors include periodontal disease, tobacco smoking and diet, and appear to trigger disease onset in genetically susceptible individuals.^2^ More recently proposed RA risk factors include the mucosal commensal microbiota. There is extensive cross-talk between the microbiota and the host, starting early in life with the development of the normal immune system; the microbiota may be implicated in the development of RA.

The gut lumen holds the vast majority of the commensal microbiota, and has intimate proximity to both the immune system via the gut associated lymphoid tissue, and the systemic circulation. A key gut microbiota taxonomic association demonstrated in RA patients, which presents early in the course of RA is an abundance of *Prevotella spp*, and particularly *Prevotella copri* (*P*.*copri*).^3,4^ Further evidence supporting the pathophysiological relevance of *P*.*copri* in RA is provided by a positive correlation with clinical parameters in RA patients.^4^ In addition to promoting disease activity, the gut microbiota may also influence patient response to treatment.^4,5^ The gut microbiota therefore represents a potential therapeutic target in RA, both for modulation of disease and for improving response to established therapeutics.

Identifying how RA genetic and environmental risk factors interact with one another may shed light on the underlying biology of disease. The influence of host genetic factors on the microbiota in RA remains relatively unexplored. An important influence is highly plausible: host genetics shape the biochemical and immune environment in which the microbiota reside, and furthermore the cumulative influence of the genetic risk loci in RA is predominantly mediated by immune pathways.^2^ Whilst there are a number of factors which influence microbiota composition, host genetic factors account for a considerable proportion of variance, with some taxa being 40% heritable.^6^

The purpose of this study was to investigate the influence of host genetic factors on the gut microbiota in RA in the absence of disease. The use of a population sample TwinsUK,^7^ whilst excluding RA affected participants allows for isolation of RA genetic factors and gets around the important issues of confounding by RA disease and its treatment - thereby achieving a human model of the genetic association with the microbiota in RA. RA polygenic risk score in unaffected TwinsUK participants was calculated and association determined with composition of the gut microbiota. Validation studies confirmed and extended our findings by examining the gut microbiota composition in first-degree relatives of RA patients (SCREEN-RA cohort) in relation to shared epitope positivity, and a pre-disease state as defined by clinical RA parameters.^8^

## Methods

### Participants: TwinsUK

Participants recruited to the study were members of the TwinsUK cohort, the largest UK registry of adult twins,^7^ with the exception that participants having a diagnosis of RA (identified by questionnaire, follow-up telephone and clinical visit confirmation) as well as their unaffected co-twin were excluded. TwinsUK participants comprised 93% females, having a median age of 63 (**Table 1**).

**Table 1.**
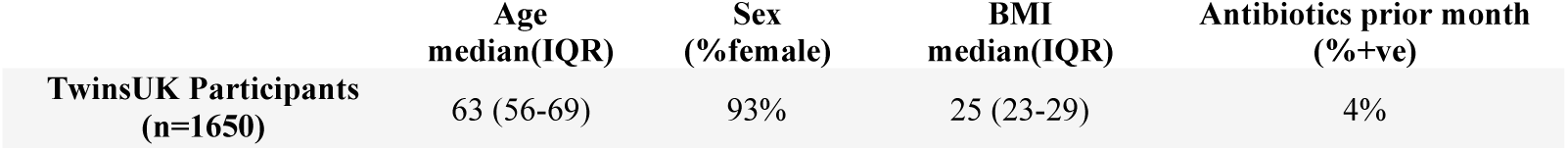
Summary of demographics for TwinsUK participants.

### Polygenic Risk Score for RA in TwinsUK

Blood samples from TwinsUK participants obtained at clinical visit were used to determine genotype using the Illumina HumanHap300 BeadChip and the Illumina HumanHap610 QuadChip. Non-genotyped variants were imputed using 1000 Genomes and Haplotype Reference Consortium (HRC) reference panels.^7^ The polygenic risk score (PRS) assigns an individual a single numerical value for risk of disease conferred by genetic factors.^9^

The NCBI database of GWAS summary statistics for RA was used to identify 233 published single nucleotide polymorphisms (SNPs) associated with RA at genome-wide significance (p=5e-8) (**Supplementary Table 1**), of which 117 had been replicated across studies (**Supplementary Table 2**).^10^ A study inclusion criterion of European ancestry ensured ethnic concordance with TwinsUK.^7,11^ The PRS was tested for RA predictive value in 6,776 participants from UK Biobank including 2,686 RA cases and 4,090 controls. Diagnosis of RA in UK Biobank participants had been determined using hospital episode statistics (HES) data supplied by NHS Digital. Logistic regression of RA cases and unselected controls against PRS, adjusting for age, sex and smoking history, was applied. Standardised coefficients are reported.

Risk allele dosage of SNPs present within TwinsUK was extracted using Plink 1.9. Of the SNPs identified in the literature 227 were available in TwinsUK. Pruning was applied to account for linkage disequilibrium ^12^ Missing allele dosages were imputed and replaced with the mean value across the respective SNPs. The risk allele dosage was multiplied by the SNP-RA association effect size, to produce a weighted PRS.^9^

High PRS individuals in our dataset did not demonstrate RA seropositivity: only 9/500 individuals in the TwinsUK sample with available serum were anti-citrullinated protein antibody (ACPA positive; defined as >5u/mL), distributed similarly between high and low genetic risk groups (high 2:93; low 7:398).

### Microbiota profiling: TwinsUK

Microbiota composition of gut (stool, n=1650) samples was assessed using the 16S rRNA marker gene, with sequencing of the V4 variable region using barcoded primers (F515/R806). Samples were processed as previously described.^6^ Briefly, faecal samples were collected during clinical visits or were posted in sealed ice packs and frozen on arrival at the lab at −80° C. Stool samples were sent as 35 batches on dry ice to Cornell University, where DNA was extracted and sequenced on an Illumina MiSeq platform.^6^

16S sequences were demultiplexed in QIIME. Amplicon sequence variants (ASVs) were generated using the DADA2 package in R^13^ using the pipeline described below, applied separately for each sequencing run (n = 35), until the final merge step.^14^ Sequences were trimmed, error estimated within the forward and reverse reads for each sample, and the ASV algorithm applied to infer the biological sequence. Forward and reverse ASVs were joined. Chimeras were removed and the total dataset merged, followed by taxonomic assignment with SILVA1.3.2.^14^ Samples having a sequencing depth of less than 10,000 reads were excluded. A phylogenetic tree was generated using the Phangorn R package. Alpha diversity was calculated on untrimmed ASV tables using four measures: Shannon index, Simpson index, observed ASVs and Faith’s phylogenetic diversity.

### Association of the gut microbiota with genetic risk of RA

Linear mixed-effects models were used to determine association between RA PRS and alpha diversity, using alpha diversity as a response variable to the RA PRS. Modelling was performed using the lme4 package in R,^15^ with fixed effect covariates age, BMI and sequencing depth and technical covariates as random effects. Standardised coefficients were reported. Differential abundance of ASVs present in more than 5% of samples at the genus level against the RA PRS was assessed using the DESeq2 R package,^16^ using fixed-effects covariates as above. Genus level refers to grouping together all ASVs with an identical genus annotation. Taxonomic assignment using the SILVA database resulted in some instances in grouping ASVs at the genus level, which are predicted to be one of a group of species. These annotations were preserved when grouping at genus level. False discovery rate (FDR) adjustment for multiple testing was applied to all models.

### Phylogenetic and community relationship within the *Prevotellaceae* Family

To investigate further the *Prevotella spp*. associations identified, the phylogenetic and community relationships between *Prevotella_7* and *Prevotella_9* were investigated. To examine community relationships, ASVs were grouped into compositional clusters, or balances. Briefly, ASVs present in at least 10 individuals were correlated and transformed as implemented in Morton et al.^17^ This creates clades in which interacting species are closer neighbours in a clade than loosely related ones. The phylogenetic tree was subsetted to *Prevotellaceae* and visualised using the Phyloseq R package.^18^

### Abundance of Prevotella spp. in RA discordant twin pairs

To be confident that the *Prevotella spp* findings in non-RA individuals with genetic risk for RA were not reflecting a “resilience-associated” microbiota (i.e. protective factors against developing disease despite having high genetic risk), we calculated the abundance of these species in 19 monozygotic RA discordant female twin pairs from TwinsUK, which had been excluded from the previous analysis because of the presence of disease. If associated with resilience we would expect the *Prevotella spp* to be preferentially expressed in the unaffected twin of each discordant pair. RA diagnosis had been confirmed during a clinical visit to a Rheumatologist or RA-trained research assistant. ASVs were generated as above and relative abundance of *Prevotella spp*. in RA discordant twin pairs calculated using the Phyloseq R package^18^ (**Figure 6**).

**Figure 1.**
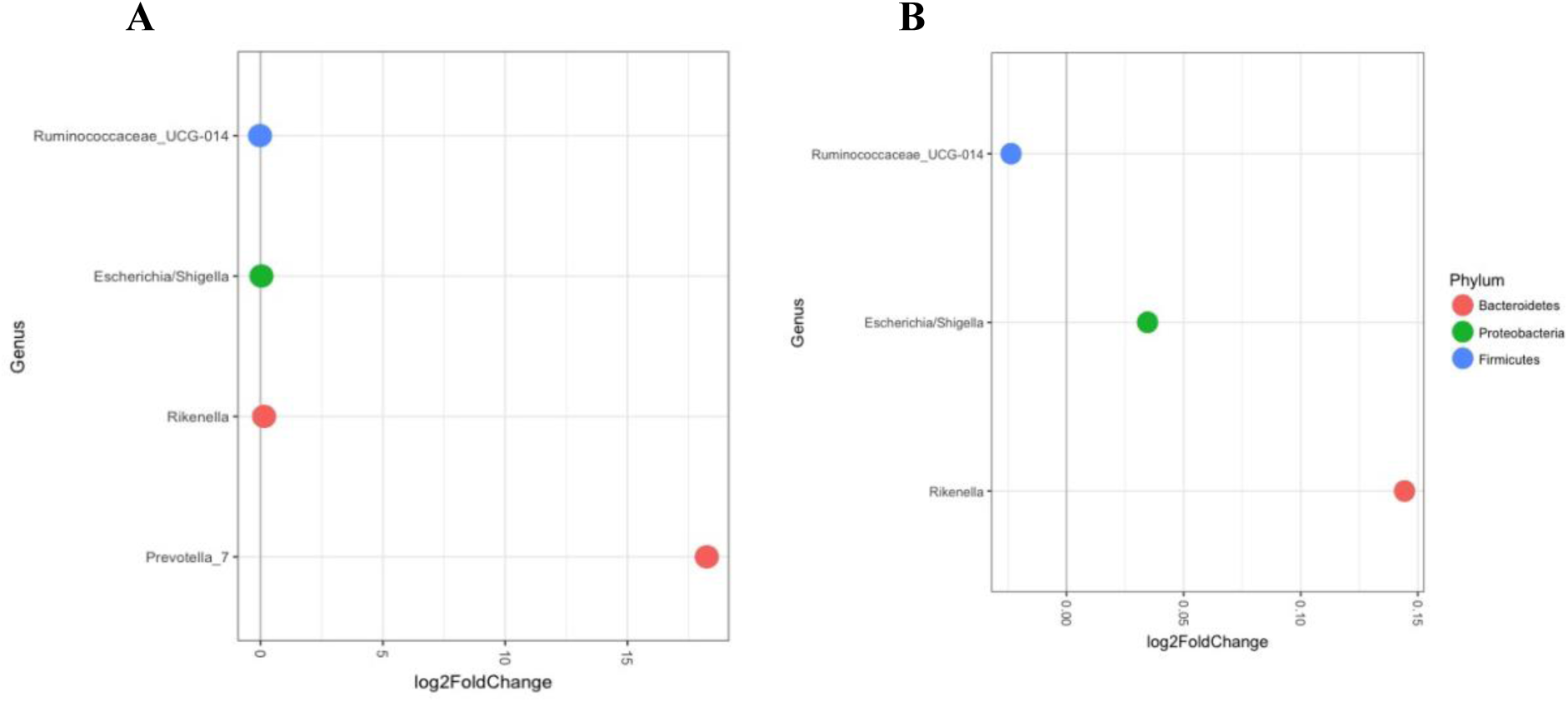
Genus level taxonomic association of the gut microbiota with PRS in TwinsUK participants. All FDR-adjusted significant results are included in **A**. Prevotella (genus) was the strongest association (p.adj <1e-7). Due to the scale and comparative difference in log-fold change in Prevotella, this taxon is excluded in **B** to allow visualisation of the 3 other associations. Other taxa associations within the gut microbiota were Ruminococcaceae (Padj = 0.0451), Rikenella (P.adj = 0.0178) and Shigella (P.adj = 0.0178).

**Figure 2.**
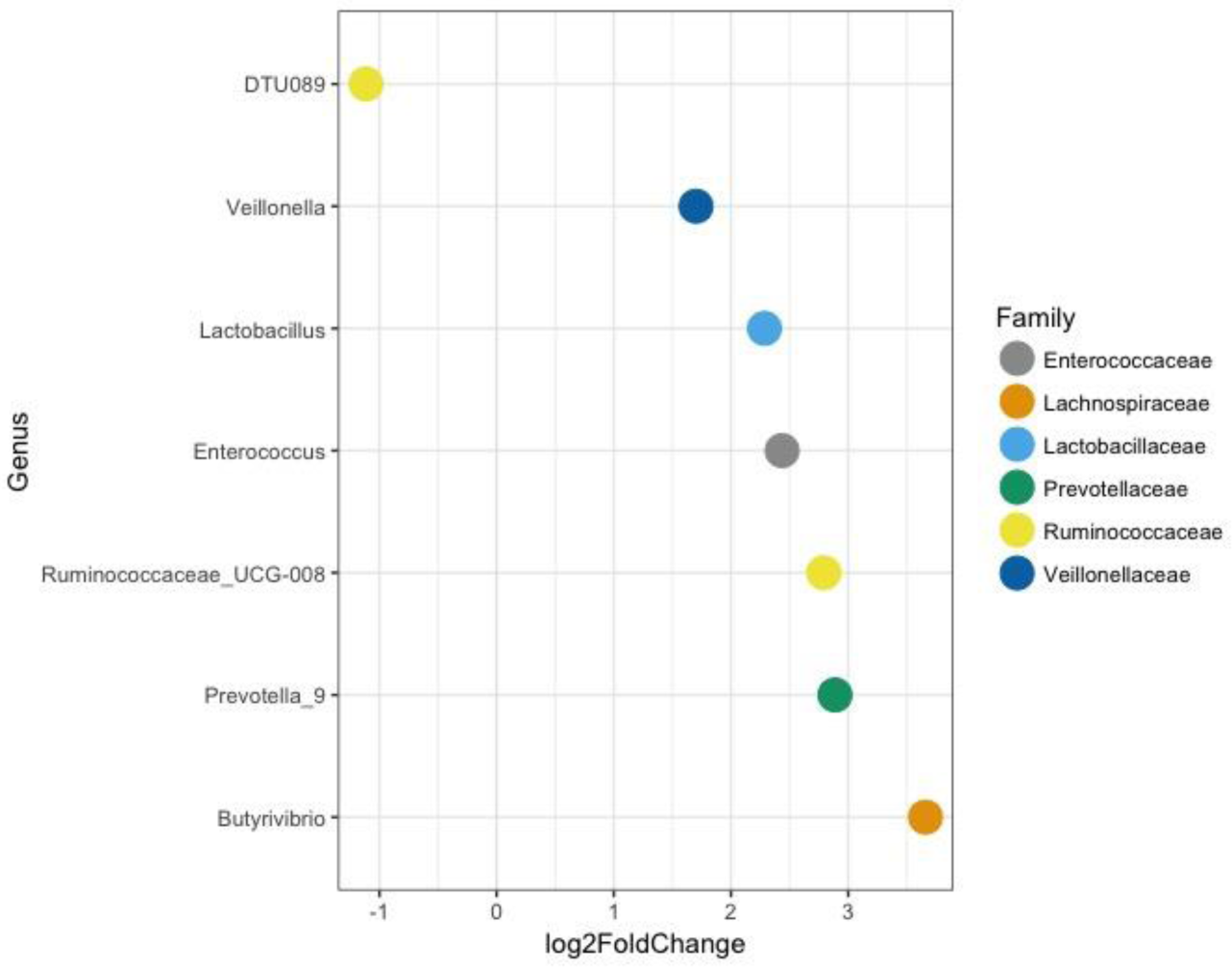
Differential abundance of the pre-clinical RA gut microbiota in the SCREEN-RA cohort, at the genus level, compared to age, sex and BMI matched controls. Prevotella (genus) was associated with pre-clinical RA (p.adj = 0.021). There were 6 additional taxa associations assigned at genus level: *Lactobacillus* (P.adj = 2.6e-4), *Butyrivibrio* (P.adj = 0.018), *Ruminococcaceae_UCG-D08* (P.adj = 0.018), *Enterococcus* (P.adj=0.018), *Ruminococcaceae DTU089* (P.adj. P = 0.042), and *Veilllonella* (P.adj = 0.042).

**Figure 3.**
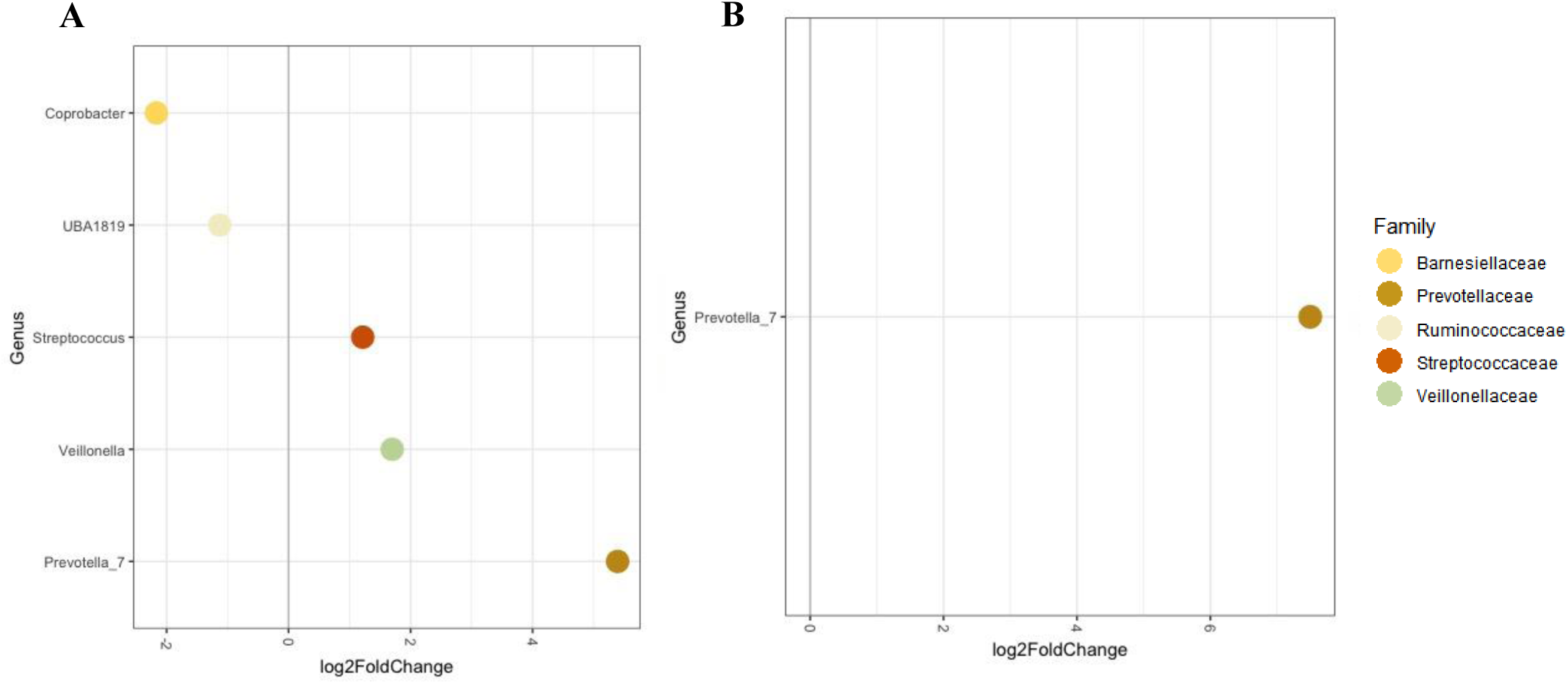
Differential abundance of the gut microbiota in SCREEN-RA participants with at least 1 copy of HLA-DRB1 shared epitope RA risk alleles, compared to age, sex and BMI matched controls. In A all participants were analysed. *Prevotella* showed the most substantial log fold change of 5 (P.adj = 3.48e-2). Other genus associations were *Veillonella* (p.adj = 3.48e-2), *Streptococcus* (P.adj = 3.48e-2), *Ruminococcaceaae UBA1819* (P.adj = 3.48e-2) and *Coprobacter* (P.adj = 2.03e-3). In **B** participants with RA associated symptoms – tender or swollen joints were excluded, here, *Prevotella* solely remained associated with shared epitope positivity (p.adj=1.12e-3).

**Figure 4.**
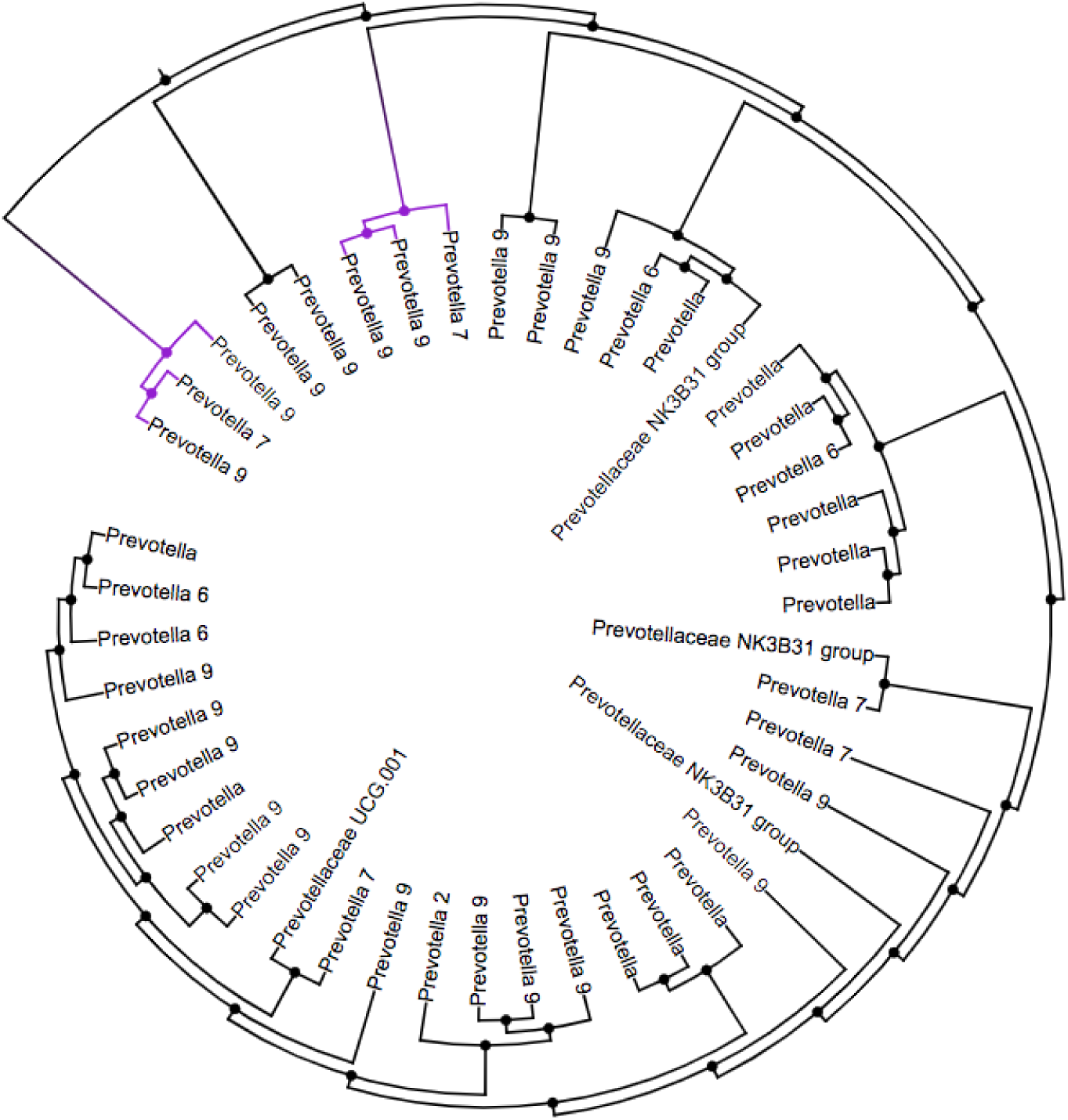
Cluster tree showing a community relationship between *Prevotella spp*. within stool samples from TwinsUK participants. *Prevotella_9* and *Prevotella_7* show community inter-relationship. *Prevotella_9* and *Prevotella_7* are the only 2-taxon clusters present, and cluster together more frequently than any other others clusters. Environmental niche similarity and biological interdependence of *Prevotella_7* and *Prevotella_9* is demonstrated.

**Figure 5.**
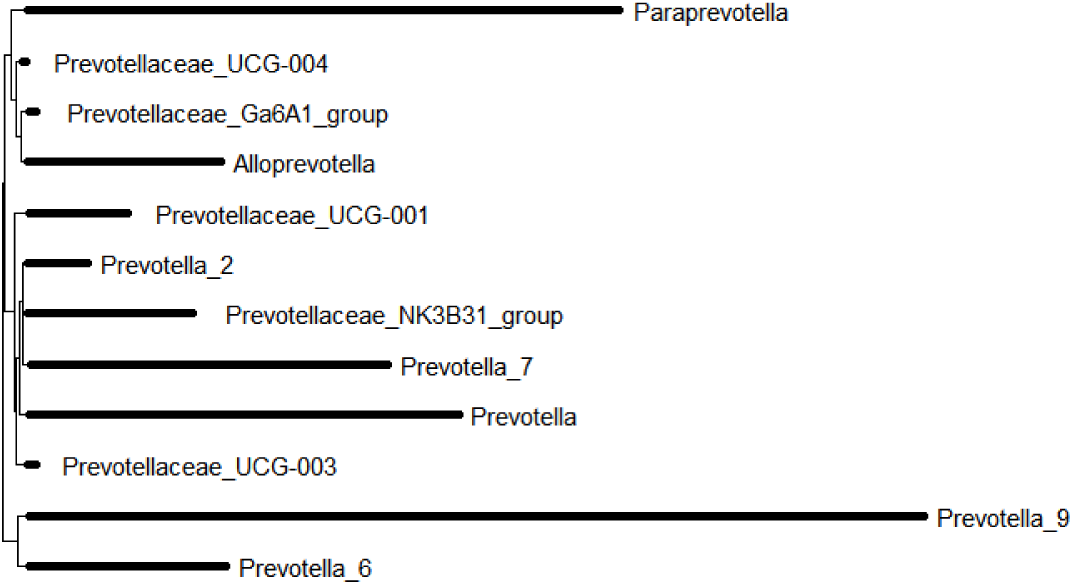
Phylogenetic tree for Prevotellaceae ASVs within TwinsUK participants. *Prevotella_9* and *Prevotella_7* are phylogenetically distinct from each other.

**Figure 6.**
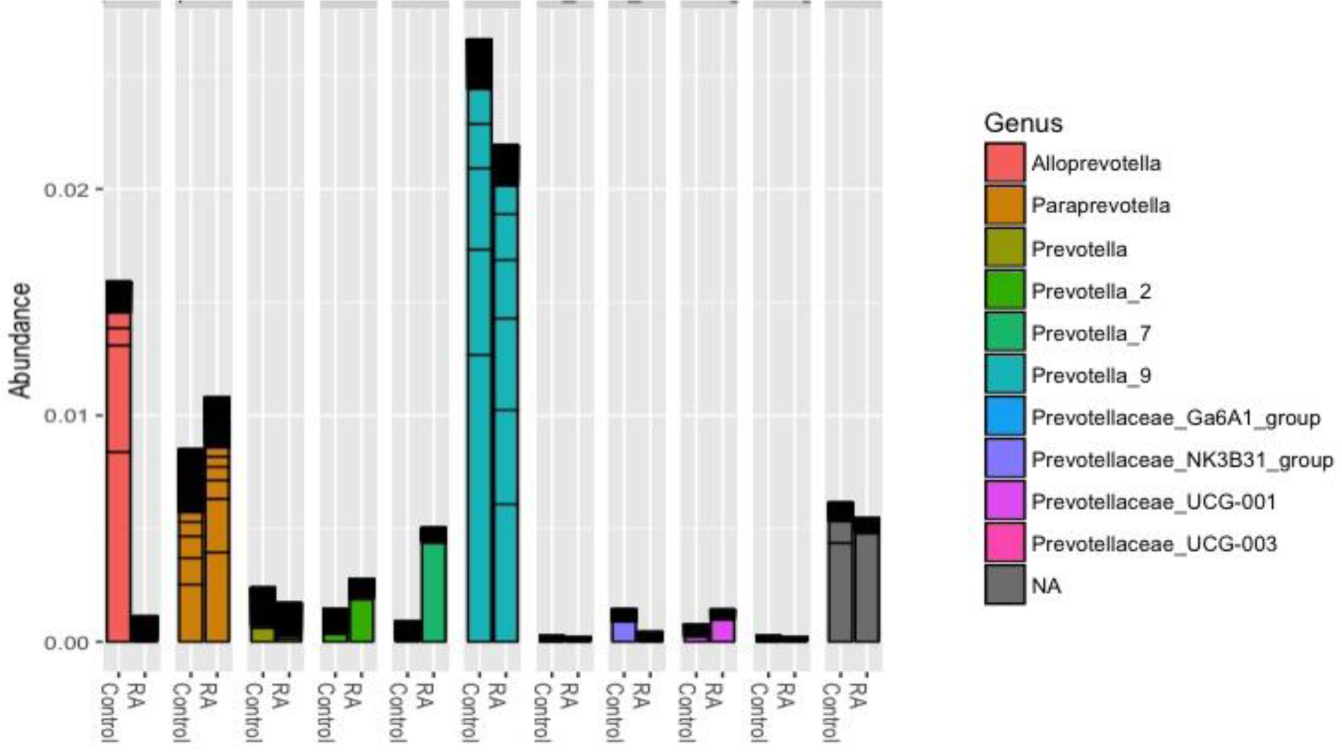
Relative abundance of *Prevotella spp*. in RA-discordant twin pairs from TwinsUK. Prevotella_7 was more abundant in RA affected co-twins, compared to non-RA co-twins.

### Participants: SCREEN-RA

To validate the results from TwinsUK, an analysis of RA genetic predisposition and microbiota association was performed in participants of SCREEN-RA, a Swiss multicentre cohort of first-degree relatives of RA patients.^8^ Within SCREEN-RA participants (n=133), pre-clinical RA cases (n = 83) were identified based on the European League Against Rheumatism terminology for pre-clinical phases of RA,^8^ and matched with 50 controls who were also first degree relatives of RA patients. Briefly, pre-clinical RA was detedefined on the basis of serum positivity for anti-citrullinated protein antibody (ACPA) or rheumatoid factor and/or symptoms and signs associated with possible RA with or without undifferentiated arthritis.^8^ Patients with a diagnosis of RA were excluded from the analysis and all participants were examined by a trained nurse. Of SCREEN-RA participants, 84% were female and median age was 57 (**Table 2**).

**Table 2.** Summary of demographics for SCREEN-RA participants.

|  | Age<br>median(IQR) | Sex<br>%female | BMI<br>median(IQR) | Swollen joints<br>median(IQR) | Tender joints<br>median(IQR) | ACPA<br>positivity<br>n(%) | RF<br>positivity<br>n(%) | SE<br>positivity<br>n(%) |
| --- | --- | --- | --- | --- | --- | --- | --- | --- |
| Pre-RA<br>n=83 | 58 (50-66) | 89 | 24 (22-27) | 1(0-3) | 1 (0-2) | 38 (46) | 28 (34) | 42(53) |
| FDR<br>controls<br>n=50 | 55 (47-62) | 78 | 24 (22-27) | 0 (0-1) | 0 (0-1) | 0 (0) | 0 (0) | 32(65) |

### Shared Epitope Genotyping: SCREEN- RA cohort

Uncoagulated blood was taken from patients and controls and stored frozen at –20° until DNA extraction. DNA was extracted using a modification of the salt-out technique (Nucleon TM, Scotlab, UK). HLA-DRB1 polymorphism was determined by reverse polymerase chain reaction - sequence-specific oligonucleotide primers hybridization and by polymerase chain reaction - sequence-specific primers using commercial reagents validated at the National Reference Laboratory for Histocompatibility. Within the DRB1*04 genotype the method discriminates all major subtypes in different allele groups. HLA-DR1, DR4 and DR14 alleles that are negative for the SE70-74 motif are also discriminated. In a second step, the SE-positive typing ambiguities were analyzed by PCR-SSP in order to determine the final 4-digit typing result.

### Microbiota Profiling and Statistical Analysis: SCREEN-RA cohort

Gut microbiota 16S rRNA data was collected as previously described.^8^ Briefly, the DNA Genotek OMNIgene·Gut Stool Microbiome Kit was used to collect, store and ship the stool samples. After sample processing and DNA extraction, the V4 region of the 16S rRNA gene was amplified using barcoded primers (F515/R806), and sequenced on an Illumina MiSeq, with ASVs generated as per the TwinsUK cohort to ensure compatibility. Differential abundance of taxa in the gut microbiota in association with pre-clinical RA and SE positivity was assessed against all genus present in greater than 5% of samples using the DESeq2 R package.^16^ Biological covariates were not required to adjust for as these factors were matched between the case and control groups.

## Results

TwinsUK participants comprised 1650 adult twin volunteers the majority of whom were female. In order to consider genetic risk factors isolated from the presence of disease, the exclusion criteria were as follows: diagnosis of RA, or twin sibling with a diagnosis of RA. **Table 1** summarises the characteristics of the sample.

### Polygenic Risk Score

The polygenic risk score was normally distributed in the TwinsUK sample with RA cases and twin siblings excluded (Shapiro-Wilk normality test P= 0.24). The score ranged from 12.6-71.3; the median score was 42.63, the interquartile range 38.89 – 48.23. Application of the PRS in 6,776 UK biobank participants, of which 2,686 had a HES diagnosis of RA, confirmed that the score is predictive of RA. The odds ratio (OR) for the PRS was modest but highly statistically significant:OR = 1.34; p = 4.17e^-8^).

### Genetic risk for RA and alpha diversity

We determined whether genetic risk of RA is associated with alpha (within sample) diversity. There was no detectable association between RA PRS and Shannon Index, Simpson Index, Observed ASVs or Faith’s phylogenetic diversity (**Table 3**).

**Table 3.** Association of genetic risk for RA (PRS) and alpha diversity of the gut microbiota in TwinsUK participants.

| Alpha diversity | Estimate | Std. Error | R<sup>2</sup> | P |
| --- | --- | --- | --- | --- |
| Observed | -0.011 | 0.021 | 0.251 | 0.6 |
| Shannon | -0.007 | 0.024 | 0.026 | 0.76 |
| Simpson | -0.023 | 0.034 | 0.006 | 0.41 |
| Faith's | -0.009 | 0.021 | 0.116 | 0.65 |

### RA PRS and the gut microbiota

The gut microbiota was assessed for association with RA PRS following a non-targeted approach at the genus level. Of all microbial taxa present in more than 5% of stool samples, *Prevotella* was the strongest association and showed an 18-fold log base 2 higher differential abundance with genetic risk for RA (P.adj < 1e-7).

### SCREEN-RA Cohort: association with pre-clinical RA and shared epitope alleles

We sought to confirm our findings in gut microbiota from the SCREEN-RA cohort by analysing the association between the main genetic risk factor for RA - HLA-DRB1 shared epitope (SE), and the intestinal microbiota. As the cohort comprises first-degree relatives of RA patients it has a higher genetic risk for RA than the general population. Participants had also been genotyped for SE risk alleles.

*Prevotella* (genus) was positively associated with pre-clinical RA (p.adj = 0.021; **Figure 2**). *Prevotella_9* is listed as potential *P*.*copri* at 97% similarity within the SILVA database.^14^ *Prevotella* (genus) was associated with HLA-DRB1 SE RA risk alleles in the SCREEN-RA cohort (n=133; p.adj = (0.0348) and more strongly so in a subgroup analysis in which the 44 participants with swollen joints were removed, in order to isolate genotype from RA pathophysiology (n=89; p.adj=1.12e-3), **Figure 3**. In the subgroup analysis of differential abundance against SE positivity in asymptomatic participants only, *Prevotella_7* was the only remaining taxon association. This finding validates the TwinsUK *Prevotella_7* association.

### Phylogenetic and community relationship within *Prevotellaceae*

As *Prevotella spp*. are the strongest taxon associations in our results and of particular interest in RA, we further investigated the phylogenetic and community relationships within *Prevotellaceae*. In particular, we were interested in the relationship between *Prevotella_7* and *Prevotella_9* as these were associated with genetic risk and pre-clinical RA, respectively. Cluster analysis demonstrated a community relationship between *Prevotella spp*. As exact matches between ASV sequences and reference species were not made, the SILVA database facilitated annotation at the species level according to groups of similar species ASV sequence.^14^ Using this database annotation at 97% similarity, it is inferred that *Prevotella_9* potentially represents *Prevotella copri*, whilst *Prevotella_7* is a different species of uncertain annotation. *Prevotella_7* and *Prevotella_9* were the only 2 species clusters within the *Prevotellaceae* family, and clustered together more frequently than any other group, suggesting an inter-dependent community relationship between these taxa (**Figure 4**). Visualisation of the *Prevotellaceae* phylogenetic tree demonstrated that *Prevotella_7* and *Prevotella_9* are phylogenetically distinct from one another (**Figure 5**).

### Abundance of Prevotella spp. in RA discordant twin pairs

We sought to investigate the potential role of *Prevotella_7* in RA. The *Prevotella_7* associations demonstrated may be implicated in RA pathogenesis, and may, indeed, be increased in RA patients. Conversely, given that the RA-unaffected TwinsUK participants are beyond the mean age of RA onset, yet have high genetic risk, these participants may be resilient to RA. Correspondingly, the genetic risk and microbiota associations demonstrated may be microbial markers of RA resilience. In this instance, the ratio of *Prevotella_7:Prevotella_9 (copri)* would be expected to be higher in unaffected twins.

The relative abundance of *Prevotella spp*. was calculated in 18 monozygotic female RA-discordant twin pairs excluded from our previous analysis. A higher abundance of *Prevotella*_7 was demonstrated in the RA affected twins compared to control twins: 0.005 and 0.001, respectively, supporting the hypothesis that *Prevotella_7* is implicated in RA aetiology. The difference in abundance however was not statistically significant. The proportion of *Prevotella_7* (species prediction uncertain) to *Prevotella_9* (predicted *Prevotella* copri) was higher in the RA affected twins than the control twins: 0.4 and 0.01, respectively, suggesting that *Prevotella_7* is not a marker of RA resilience.

## Discussion

This study demonstrates a link between host genetic risk for RA and the gut microbiota in a large human cohort. Gut microbiota associations have been demonstrated in the absence of clinically detectable disease because participants with RA, along with their unaffected co-twins, were removed from the study. The strongest association with genetic risk for RA in the gut microbiota was an increase in abundance of *Prevotella_7* (18-fold log-change; p.adj<1e-7), and the *Prevotella_7* association was confirmed by our analysis of the SCREEN-RA cohort in which this taxon was positively correlated with shared epitope positivity (7-fold log change; p.adj=1.12e-3). *Prevotella_7* was demonsrated to share a biological interdependence and niche similarity with *Prevotella_9 (copri)*.

There is considerable interest in *Prevotella copri* as a potential mediator of RA pathology, and it is a candidate keystone taxon enriched in the gut microbiota of newly diagnosed RA patients. Since this observation was made, the human immune response to this microbe has been of considerable interest. Antibodies to *P*.*copri* have been shown to associate with disease severity and innate- and Th1 and Th17 immune responses in RA^4,19^.

Functional work has suggested a role for *P*.*copri* in Th17 cell differentiation.^20^ That the abundance of *P*.*copri* abates after treatment with DMARDS, as shown in a longitudinal study, ^3^ and is also associated with other inflammatory conditions,^21^ has fuelled speculation that inflammation is a pre-requisite for *P*.*copri* proliferation within the gut, relative to other taxa.^19,22^ *P*.*copri* may have adapted to thrive in a pro-inflammatory environment, and may further promote the inflammatory milieu, thereby enhancing its own favoured environmental niche.^22^ In doing so, *P*.*copri* is suggested to contribute to RA pathology. According to this model, a human–microbiota interspecies positive feedback loop is proposed. Another example of such a model is the hijacking of complement cascade by *Porphyromonas gingivalis*.^23^

An increase in abundance of three other taxa - *Ruminococaceae, Rikenella* and *Shigella* - in association with RA PRS was also observed. *Ruminococcaceae* and *Rikenella* have been shown previously to be in higher abundance in RA patients compared to controls.^3,24^ The evidence for a role of these taxa in RA is much weaker than for *P*.*copri* - with lower replication across studies, and no functional link reported to date. However, the association both with RA patients and with genetic risk for RA in a large, non-disease cohort is interesting, and merits further investigation.

Whilst causality is not determinable from a cross sectional association study, our results provide robust evidence to indicate that genetic factors in RA may regulate the abundance of *Prevotella* seen in the gut microbiota of people with RA. Both TwinsUK and SCREEN RA cohorts were balanced for ACPA positivity in relation to RA genetic risk loci. A previous study of new onset RA patients and healthy participants reported an inverse association of HLA genotype with abundance of *Prevotella copri*, in the opposite direction to that expected.^4,25^ This study used operational taxonomic units (OTUs) and potential confounding factors were not examined; it is not possible accurately to determine species from OTUs. The present study using ASVs in a very large sample provides more substantial evidence for an association of genotype with the RA gut microbiota.

The findings suggest that the microbiota is altered prior to disease and are in accord with previous reports of *Prevotella spp*. in pre-RA^8^ and RA. Indeed, we find similar in cases of pre-clinical RA in first degree relatives of RA patients. In a recent study of the SCREEN-RA cohort an enrichment of *Prevotellaceae* in the gut microbiota in pre-RA was shown. In the study by Alpizaar-Rodriguez *et al*.^8^ no statistically significant *Prevotellaceae* genera were shown to drive the association. Therefore in the present study data were re-analysed using the more recent method of ASVs. ASVs offer an updated alternative to traditional clustering based methods generating OTUs from 16S sequence. As opposed to grouping sequences based on a similarity threshold as for OTUs, it is now possible to use the error rate to infer the original biological sequence, and produce units of matched sequence. ASVs offer higher resolution, therefore, and have greater biological relevance.^26^ The ASV analysis showed an FDR-adjusted significant *Prevotella* genus association with a species level group annotation indicating potential *Prevotella copri*. Our follow-up study of pre-RA further revealed six novel genus associations which had not been evident in the original OTUs– *Lactobacillus, Enterococcus, Faecalibacterium, Ruminococcaceae, Veilllonella* and *Butyrivibrio*. Of the six associations, the first four have been reported associated with RA previously,^3,27,28^ and *Ruminococcaceae* was seen associated with RA PRS in TwinsUK.

There are a number of limitations to the study. First, in the pre-clinical RA follow-up analysis both cases and controls were first degree relatives of RA patients, with increased genetic risk compared to the general population. It is possible that the pre-clinical RA cases had higher genetic risk than the controls, but as full genotyping was not available, we were unable to confirm this. The TwinsUK cohort is predominantly female – 93% of participants in our study were female. The SCREEN-RA cohort is slightly more balanced in terms of sex, however population prevalence of RA is much higher in females than males. As the age of onset of RA is 30-65,^29^ the TwinsUK participants are less likely to develop disease than the SCREEN-RA participants. However, this is of benefit to the design of our study of genetically high risk yet unaffected individuals and helps us to understand microbiota differences in the absence of disease.

On a technical note, this work highlights the value of using updated methods of amplicon sequence variants – ASVs - which can detect taxonomic variation overlooked by OTU based methods: there are likely to be further microbiota associations with RA as yet undetected due to limitations in 16S methods. Annotation of ASVs with the SILVA database demonstrates differences at the species level which would be overlooked using other reference databases. Finally, modelling of the community relationship provides valuable insight into the underlying biology. Future studies should take advantage of these methods.

Taken together, these results support the hypothesis that microbiota is altered in individuals with genetic predisposition to RA before the onset even of pre-clinical RA. The work sheds light on our understanding of microbiota in RA and addresses the cause vs consequence issue; – if microbial alteration precedes disease, the microbiota may lie on the causal pathway. However we cannot yet exclude the possibility that *Prevotella spp*. are bystanders. Additionally, having RA genetic risk and *Prevotella spp*. is not sufficient for disease development, but rather may be one of several “hits” required for progression to RA. The identification of pre-clinical RA represents an important clinical target in early disease intervention and is currently the subject of multiple immune-modulating clinical trials. The microbiota may offer the opportunity for modulation of pre-disease pathways.

## Conclusions

Gut microbiota abundance is associated with RA risk genotype even in the absence of disease. Genotype may mediate key taxonomic associations of the gut microbiota with RA, particularly *Prevotella spp*.^3,4,30^ suggesting that theses species play a role very early in development of RA.

## Data Availability

Data is available from TwinsUK.

## Acknowledgements and affiliations

This work was funded by Arthritis Research UK Special Strategic Award (grant 21227). Twins UK receives funding from the Wellcome Trust; the National Institute for Health Research (NIHR) Clinical Research Facility at Guy’s & St Thomas’ NHS Foundation Trust and NIHR Biomedical Research Centre based at Guy’s and St Thomas’ NHS Foundation Trust and King’s College London. Claire J. Steves is funded by the Wellcome Trust (grant WT081878MA). Philippa Wells is supported by a joint PhD studentship from the Medical Research Council (MRC) and Clinical Disease Reasearch Foundation (CDRF).

The authors would like thank the participants of the TwinsUK cohort.

## Author Contributions

CS, FW, BK and AC conceived the project. PW developed the theory, performed the experiments, analysed and interpreted the data, and took the lead in writing the manuscript. CS and FW supervised the analysis. CS encouraged development of the work through leading collaboration with the SCREEN-RA cohort, and inclusion of investigation of community structure, and abundance of taxa in RA discordant twins. AA performed analysis of the community structure. FW, PW and CS collected data from TwinsUK. MF assisted with UKBiobank genotype data. All authors provided ctitical feedback and contributed to the final manuscript.

## Supplementary Tables

**Supplementary Table 1.** SNPs included in the RA PRS.

| CHR | SNP | A1 | A2 |
| --- | --- | --- | --- |
| 1 | rs624988 | T | C |
| 1 | rs840016 | C | T |
| 1 | rs923880 | C | T |
| 1 | rs72717009 | T | C |
| 1 | rs72717009 | T | C |
| 1 | rs2317230 | T | G |
| 1 | rs6593803 | T | C |
| 1 | rs2228145 | A | C |
| 1 | rs2228145 | A | C |
| 1 | rs12565755 | C | T |
| 1 | rs2105325 | C | A |
| 1 | rs2105325 | C | A |
| 1 | rs12140275 | A | T |
| 1 | rs12140275 | A | T |
| 1 | rs3890745 | T | C |
| 1 | rs28411352 | T | C |
| 1 | rs28411352 | T | C |
| 1 | rs2301888 | G | A |
| 1 | rs2301888 | G | A |
| 1 | rs2301888 | G | A |
| 1 | rs12131057 | G | A |
| 1 | rs2476601 | A | G |
| 1 | rs6679677 | A | C |
| 1 | rs2476601 | A | G |
| 1 | rs2476601 | A | G |
| 1 | rs2476601 | A | G |
| 1 | rs6679677 | A | C |
| 1 | rs2476601 | A | G |
| 1 | rs2476601 | A | G |
| 1 | rs2476601 | A | G |
| 1 | rs11121380 | C | A |
| 1 | rs3890745 | T | C |
| 1 | rs3890745 | C | T |
| 1 | rs998731 | C | T |
| 2 | rs6732565 | A | G |
| 2 | rs6732565 | A | A |
| 2 | rs11676922 | T | A |
| 2 | rs10865035 | A | G |
| 2 | rs11676922 | T | A |
| 2 | rs9653442 | C | T |
| 2 | rs9653442 | C | T |
| 2 | rs11900673 | T | C |
| 2 | rs13385025 | A | G |
| 2 | rs6715284 | G | C |
| 2 | rs6715284 | G | C |
| 2 | rs1980422 | C | T |
| 2 | rs1980422 | C | T |
| 2 | rs3087243 | G | A |
| 2 | rs231735 | G | T |
| 2 | rs3087243 | G | A |
| 2 | rs3087243 | G | A |
| 2 | rs12617656 | T | C |
| 2 | rs12617656 | T | C |
| 2 | rs41005 | T | C |
| 2 | rs1406428 | T | C |
| 2 | rs10175798 | A | G |
| 2 | rs10175798 | A | G |
| 2 | rs13031237 | T | G |
| 2 | rs13017599 | A | G |
| 2 | rs34695944 | C | T |
| 2 | rs34695944 | C | T |
| 2 | rs4305317 | T | A |
| 2 | rs13015080 | A | G |
| 2 | rs13015080 | A | T |
| 2 | rs934734 | G | A |
| 2 | rs934734 | G | A |
| 2 | rs1858037 | T | A |
| 2 | rs1858037 | T | A |
| 2 | rs1858037 | T | A |
| 2 | rs7574865 | T | G |
| 2 | rs7574865 | T | G |
| 2 | rs11889341 | T | C |
| 2 | rs11889341 | T | C |
| 2 | rs11889341 | T | C |
| 3 | rs73081554 | T | C |
| 3 | rs2062583 | G | T |
| 3 | rs9860428 | C | A |
| 3 | rs9860428 | C | A |
| 3 | rs3806624 | G | A |
| 3 | rs3806624 | G | A |
| 3 | rs9826828 | A | G |
| 3 | rs6774280 | C | T |
| 3 | rs4452313 | T | A |
| 3 | rs4452313 | T | A |
| 3 | rs7624766 | G | A |
| 3 | rs13315591 | C | T |
| 3 | rs1809529 | C | T |
| 4 | rs2867461 | A | G |
| 4 | rs11933540 | C | T |
| 4 | rs13142500 | C | T |
| 4 | rs13142500 | C | T |
| 4 | rs13119723 | A | G |
| 4 | rs45475795 | G | A |
| 4 | rs11937061 | T | G |
| 4 | rs3816587 | C | T |
| 4 | rs874040 | C | G |
| 4 | rs6448432 | A | G |
| 4 | rs13137105 | G | A |
| 4 | rs2664035 | A | G |
| 4 | rs2664035 | A | G |
| 5 | rs7731626 | G | A |
| 5 | rs255758 | C | A |
| 5 | rs26232 | C | T |
| 5 | rs2561477 | G | A |
| 5 | rs2561477 | G | A |
| 5 | rs1498103 | A | T |
| 5 | rs12519788 | G | A |
| 5 | rs12519788 | G | A |
| 5 | rs657075 | A | G |
| 5 | rs657075 | A | G |
| 5 | rs657075 | A | G |
| 5 | rs6859219 | C | A |
| 5 | rs12109285 | C | T |
| 5 | rs1502644 | C | A |
| 5 | rs4336372 | C | T |
| 5 | rs4336372 | C | T |
| 5 | rs4921283 | A | G |
| 6 | rs805297 | A | C |
| 6 | rs6568431 | A | C |
| 6 | rs9372120 | G | T |
| 6 | rs9372120 | G | T |
| 6 | rs3093024 | A | G |
| 6 | rs3093023 | A | G |
| 6 | rs3093023 | A | G |
| 6 | rs1571878 | C | T |
| 6 | rs1571878 | C | T |
| 6 | rs1571878 | C | T |
| 6 | rs1854853 | A | G |
| 6 | rs1854853 | A | G |
| 6 | rs12529514 | C | T |
| 6 | rs3763309 | A | C |
| 6 | rs6457617 | T | C |
| 6 | rs9275406 | T | G |
| 6 | rs12525220 | A | G |
| 6 | rs12525220 | A | G |
| 6 | rs12525220 | A | G |
| 6 | rs660895 | G | A |
| 6 | rs7765379 | G | T |
| 6 | rs13192471 | G | A |
| 6 | rs6457620 | G | C |
| 6 | rs6910071 | G | A |
| 6 | rs9268839 | G | A |
| 6 | rs9268839 | G | A |
| 6 | rs9268839 | G | A |
| 6 | rs9269234 | G | C |
| 6 | rs9269234 | G | C |
| 6 | rs9271348 | G | A |
| 6 | rs9378815 | C | G |
| 6 | rs9378815 | C | G |
| 6 | rs284515 | G | A |
| 6 | rs284511 | C | T |
| 6 | rs7748270 | T | C |
| 6 | rs7748270 | T | C |
| 6 | rs6457617 | T | C |
| 6 | rs2596565 | A | G |
| 6 | rs2233434 | G | A |
| 6 | rs2233434 | G | A |
| 6 | rs2233424 | T | C |
| 6 | rs2233424 | T | C |
| 6 | rs2233424 | T | C |
| 6 | rs9373594 | T | C |
| 6 | rs9373594 | T | C |
| 6 | rs2451258 | T | C |
| 6 | rs2451258 | T | C |
| 6 | rs6920220 | A | G |
| 6 | rs6920220 | A | G |
| 6 | rs5029924 | T | C |
| 6 | rs7752903 | G | T |
| 6 | rs7752903 | G | T |
| 6 | rs7752903 | G | T |
| 6 | rs10499194 | C | T |
| 6 | rs6920220 | A | G |
| 6 | rs2230926 | C | A |
| 6 | rs6920220 | A | G |
| 6 | rs75908454 | C | T |
| 7 | rs42041 | G | C |
| 7 | rs4272 | G | A |
| 7 | rs4272 | G | A |
| 7 | rs6956740 | T | C |
| 7 | rs11761231 | T | C |
| 7 | rs10488631 | C | T |
| 7 | rs3807306 | T | G |
| 7 | rs67250450 | T | C |
| 7 | rs67250450 | T | C |
| 7 | rs12531711 | G | A |
| 8 | rs1600249 | T | A |
| 8 | rs2736340 | T | C |
| 8 | rs2736340 | T | C |
| 8 | rs2736337 | C | T |
| 8 | rs2736337 | C | T |
| 8 | rs2736337 | C | T |
| 8 | rs678347 | G | A |
| 8 | rs678347 | G | A |
| 8 | rs16938910 | T | C |
| 8 | rs800586 | A | G |
| 8 | rs1516971 | T | C |
| 8 | rs1516971 | T | T |
| 8 | rs998731 | T | C |
| 8 | rs998731 | T | C |
| 9 | rs11574914 | A | G |
| 9 | rs11574914 | A | G |
| 9 | rs2812378 | G | A |
| 9 | rs951005 | A | G |
| 9 | rs12379034 | G | A |
| 9 | rs12379034 | G | A |
| 9 | rs1329568 | T | G |
| 9 | rs9299346 | A | G |
| 9 | rs2072438 | T | C |
| 9 | rs3761847 | G | A |
| 9 | rs3761847 | G | A |
| 9 | rs10985070 | C | A |
| 9 | rs10985070 | C | A |
| 10 | rs10821944 | G | T |
| 10 | rs71508903 | T | C |
| 10 | rs71508903 | T | C |
| 10 | rs3824660 | C | T |
| 10 | rs3824660 | C | T |
| 10 | rs1679568 | A | G |
| 10 | rs1679568 | A | G |
| 10 | rs706778 | T | C |
| 10 | rs2104286 | C | T |
| 10 | rs706778 | T | C |
| 10 | rs706778 | T | C |
| 10 | rs12413578 | C | T |
| 10 | rs12413578 | C | T |
| 10 | rs16906916 | C | A |
| 10 | rs12570744 | T | C |
| 10 | rs4750316 | G | C |
| 10 | rs4750316 | G | C |
| 10 | rs947474 | A | G |
| 10 | rs947474 | A | G |
| 10 | rs3125734 | T | C |
| 10 | rs6479800 | C | G |
| 10 | rs6479800 | C | G |
| 10 | rs726288 | T | C |
| 10 | rs2671692 | A | G |
| 10 | rs793108 | T | C |
| 10 | rs793108 | T | C |
| 11 | rs508970 | A | G |
| 11 | rs4409785 | C | T |
| 11 | rs4409785 | C | T |
| 11 | rs10790268 | G | A |
| 11 | rs10790268 | G | A |
| 11 | rs73013527 | C | T |
| 11 | rs73013527 | C | T |
| 11 | rs73013527 | C | T |
| 11 | rs4937362 | T | C |
| 11 | rs968567 | C | T |
| 11 | rs518167 | A | G |
| 11 | rs7940423 | A | G |
| 11 | rs72991 | C | T |
| 11 | rs1079467 | C | NA |
| 11 | rs3781913 | T | G |
| 11 | rs331463 | T | A |
| 11 | rs331463 | T | A |
| 12 | rs773125 | A | G |
| 12 | rs773125 | A | G |
| 12 | rs1633360 | T | C |
| 12 | rs1633360 | T | C |
| 12 | rs11051970 | T | G |
| 12 | rs1678542 | C | G |
| 12 | rs1678542 | C | G |
| 12 | rs11045392 | C | T |
| 12 | rs3184504 | T | C |
| 12 | rs10774624 | G | A |
| 12 | rs3794271 | G | A |
| 12 | rs12831974 | C | T |
| 13 | rs9557321 | C | T |
| 13 | rs9603612 | G | C |
| 13 | rs9603616 | C | T |
| 13 | rs9603616 | C | T |
| 13 | rs4942242 | T | C |
| 13 | rs9604529 | G | A |
| 13 | rs3790022 | A | G |
| 14 | rs2582532 | C | T |
| 14 | rs2582532 | C | T |
| 14 | rs7155603 | G | A |
| 14 | rs3783637 | C | T |
| 14 | rs17118552 | G | A |
| 14 | rs1624005 | G | A |
| 14 | rs7141276 | A | G |
| 14 | rs2841277 | T | C |
| 14 | rs1957895 | G | T |
| 14 | rs3783782 | A | G |
| 14 | rs3783782 | A | G |
| 14 | rs1950897 | T | C |
| 14 | rs1950897 | T | C |
| 15 | rs1914816 | A | G |
| 15 | rs1898036 | C | T |
| 15 | rs7164176 | G | A |
| 15 | rs17374222 | A | C |
| 15 | rs8026898 | A | G |
| 15 | rs8026898 | A | G |
| 15 | rs12901682 | A | C |
| 15 | rs8032939 | C | T |
| 15 | rs8032939 | C | T |
| 15 | rs8032939 | C | T |
| 15 | rs6496667 | A | C |
| 16 | rs6500395 | C | T |
| 16 | rs2280381 | T | C |
| 16 | rs13330176 | A | T |
| 16 | rs13330176 | A | T |
| 16 | rs16973500 | T | NA |
| 16 | rs7404928 | T | C |
| 16 | rs4780401 | T | G |
| 16 | rs4780401 | T | G |
| 17 | rs72634030 | A | C |
| 17 | rs2872507 | A | G |
| 17 | rs1877030 | C | T |
| 17 | rs12601925 | G | A |
| 17 | rs12601925 | G | A |
| 18 | rs2469434 | C | T |
| 18 | rs2469434 | C | T |
| 18 | rs16977065 | T | C |
| 18 | rs2847297 | G | A |
| 18 | rs8083786 | G | A |
| 18 | rs8083786 | G | A |
| 18 | rs8083786 | G | A |
| 18 | rs6506569 | T | C |
| 18 | rs2002842 | A | C |
| 18 | rs1943199 | T | G |
| 19 | rs77331626 | G | A |
| 19 | rs7258015 | C | T |
| 19 | rs1273516 | A | G |
| 19 | rs1543922 | T | C |
| 19 | rs34536443 | G | C |
| 20 | rs4810485 | G | T |
| 20 | rs4239702 | C | T |
| 20 | rs4239702 | C | T |
| 20 | rs6138892 | C | T |
| 20 | rs6026990 | A | T |
| 21 | rs2075876 | A | G |
| 21 | rs73194058 | C | A |
| 21 | rs73194058 | C | A |
| 21 | rs2837960 | G | T |
| 21 | rs8133843 | A | G |
| 21 | rs8133843 | A | G |
| 21 | rs11203203 | A | G |
| 21 | rs1893592 | A | C |
| 22 | rs1043099 | C | G |
| 22 | rs3218251 | A | T |
| 22 | rs743777 | G | A |
| 22 | rs909685 | A | T |
| 22 | rs909685 | A | T |
| 22 | rs909685 | A | T |
| 22 | rs2283790 | G | A |
| 22 | rs11089637 | C | T |
| 22 | rs11089637 | C | T |
| X | rs5987194 | C | G |
| X | rs5987194 | C | G |

**Supplementary Table 2:** Replicated SNPs.

| CHR | SNP | A1 | A2 |
| --- | --- | --- | --- |
| 1 | rs72717009 | T | C |
| 1 | rs2228145 | A | C |
| 1 | rs2105325 | C | A |
| 1 | rs12140275 | A | T |
| 1 | rs28411352 | T | C |
| 1 | rs2301888 | G | A |
| 1 | rs2301888 | G | A |
| 1 | rs2476601 | A | G |
| 1 | rs2476601 | A | G |
| 1 | rs2476601 | A | G |
| 1 | rs6679677 | A | C |
| 1 | rs2476601 | A | G |
| 1 | rs2476601 | A | G |
| 1 | rs2476601 | A | G |
| 1 | rs3890745 | T | C |
| 1 | rs3890745 | C | T |
| 2 | rs6732565 | A | A |
| 2 | rs11676922 | T | A |
| 2 | rs9653442 | C | T |
| 2 | rs6715284 | G | C |
| 2 | rs1980422 | C | T |
| 2 | rs3087243 | G | A |
| 2 | rs3087243 | G | A |
| 2 | rs12617656 | T | C |
| 2 | rs10175798 | A | G |
| 2 | rs34695944 | C | T |
| 2 | rs13015080 | A | T |
| 2 | rs934734 | G | A |
| 2 | rs1858037 | T | A |
| 2 | rs1858037 | T | A |
| 2 | rs7574865 | T | G |
| 2 | rs11889341 | T | C |
| 2 | rs11889341 | T | C |
| 3 | rs9860428 | C | A |
| 3 | rs3806624 | G | A |
| 3 | rs4452313 | T | A |
| 4 | rs13142500 | C | T |
| 4 | rs2664035 | A | G |
| 5 | rs2561477 | G | A |
| 5 | rs12519788 | G | A |
| 5 | rs657075 | A | G |
| 5 | rs657075 | A | G |
| 5 | rs4336372 | C | T |
| 6 | rs9372120 | G | T |
| 6 | rs3093023 | A | G |
| 6 | rs1571878 | C | T |
| 6 | rs1571878 | C | T |
| 6 | rs1854853 | A | G |
| 6 | rs12525220 | A | G |
| 6 | rs12525220 | A | G |
| 6 | rs9268839 | G | A |
| 6 | rs9268839 | G | A |
| 6 | rs9269234 | G | C |
| 6 | rs9378815 | C | G |
| 6 | rs7748270 | T | C |
| 6 | rs6457617 | T | C |
| 6 | rs2233434 | G | A |
| 6 | rs2233424 | T | C |
| 6 | rs2233424 | T | C |
| 6 | rs9373594 | T | C |
| 6 | rs2451258 | T | C |
| 6 | rs6920220 | A | G |
| 6 | rs7752903 | G | T |
| 6 | rs7752903 | G | T |
| 6 | rs6920220 | A | G |
| 6 | rs6920220 | A | G |
| 7 | rs4272 | G | A |
| 7 | rs67250450 | T | C |
| 8 | rs2736340 | T | C |
| 8 | rs2736337 | C | T |
| 8 | rs2736337 | C | T |
| 8 | rs678347 | G | A |
| 8 | rs1516971 | T | T |
| 8 | rs998731 | T | C |
| 8 | rs998731 | T | C |
| 9 | rs11574914 | A | G |
| 9 | rs12379034 | G | A |
| 9 | rs3761847 | G | A |
| 9 | rs10985070 | C | A |
| 10 | rs71508903 | T | C |
| 10 | rs3824660 | C | T |
| 10 | rs1679568 | A | G |
| 10 | rs706778 | T | C |
| 10 | rs706778 | T | C |
| 10 | rs12413578 | C | T |
| 10 | rs4750316 | G | C |
| 10 | rs947474 | A | G |
| 10 | rs6479800 | C | G |
| 10 | rs793108 | T | C |
| 11 | rs4409785 | C | T |
| 11 | rs10790268 | G | A |
| 11 | rs73013527 | C | T |
| 11 | rs73013527 | C | T |
| 11 | rs331463 | T | A |
| 12 | rs773125 | A | G |
| 12 | rs1633360 | T | C |
| 12 | rs1678542 | C | G |
| 13 | rs9603616 | C | T |
| 14 | rs2582532 | C | T |
| 14 | rs3783782 | A | G |
| 14 | rs1950897 | T | C |
| 15 | rs8026898 | A | G |
| 15 | rs8032939 | C | T |
| 15 | rs8032939 | C | T |
| 16 | rs13330176 | A | T |
| 16 | rs4780401 | T | G |
| 17 | rs12601925 | G | A |
| 18 | rs2469434 | C | T |
| 18 | rs8083786 | G | A |
| 18 | rs8083786 | G | A |
| 20 | rs4239702 | C | T |
| 21 | rs73194058 | C | A |
| 21 | rs8133843 | A | G |
| 22 | rs909685 | A | T |
| 22 | rs909685 | A | T |
| 22 | rs11089637 | C | T |
| X | rs5987194 | C | G |

## References

1 MacGregor AJ, Snieder H, Rigby AS, et al. Characterizing the quantitative genetic contribution to rheumatoid arthritis using data from twins. Arthritis Rheum 2000; 43: 30–7.

2 Wells PM, Williams FMK, Matey-Hernandez ML, Menni C, Steves CJ. ‘RA and the microbiome: do host genetic factors provide the link? J Autoimmun 2019; published online March 5. DOI:10.1016/j.jaut.2019.02.004.

3 Zhang X, Zhang D, Jia H, et al. The oral and gut microbiomes are perturbed in rheumatoid arthritis and partly normalized after treatment. Nat Med 2015; 21: 895–905.

4 Scher JU, Sczesnak A, Longman RS, et al. Expansion of intestinal Prevotella copri correlates with enhanced susceptibility to arthritis. eLife 2013; 2: e01202.

5 Isaac S, Artacho A, Nayak R, et al. Op0119 the Pre-Treatment Gut Microbiome Predicts Early Response to Rheumatoid Arthritis Therapy. Ann Rheum Dis 2019; 78: 133–4.

6 Goodrich JK, Waters JL, Poole AC, et al. Human Genetics Shape the Gut Microbiome. Cell 2014; 159: 789–99.

7 Verdi S, Abbasian G, Bowyer RCE, et al. TwinsUK: The UK Adult Twin Registry Update. Twin Res Hum Genet Off J Int Soc Twin Stud 2019;: 1–7.

8 Alpizar-Rodriguez D, Lesker TR, Gronow A, et al. Prevotella copri in individuals at risk for rheumatoid arthritis. Ann Rheum Dis 2019; 78: 590–3.

9 Khera AV, Chaffin M, Aragam KG, et al. Genome-wide polygenic scores for common diseases identify individuals with risk equivalent to monogenic mutations. Nat Genet 2018; 50: 1219–24.

10 Jones HJ, Hubbard L, Mitchell RE, et al. Association of Genetic Risk for Rheumatoid Arthritis With Cognitive and Psychiatric Phenotypes Across Childhood and Adolescence. JAMA Netw Open 2019; 2. DOI:10.1001/jamanetworkopen.2019.6118.

11 Duncan L, Shen H, Gelaye B, et al. Analysis of polygenic risk score usage and performance in diverse human populations. Nat Commun 2019; 10: 1–9.

12 Vilhjálmsson BJ, Yang J, Finucane HK, et al. Modeling Linkage Disequilibrium Increases Accuracy of Polygenic Risk Scores. Am J Hum Genet 2015; 97: 576–92.

13 Callahan BJ, McMurdie PJ, Rosen MJ, Han AW, Johnson AJA, Holmes SP. DADA2: High resolution sample inference from Illumina amplicon data. Nat Methods 2016; 13: 581–3.

14 Callahan B. Silva taxonomic training data formatted for DADA2 (Silva version 132). 2018; published online Feb 13. DOI:10.5281/zenodo.1172783.

15 Bates D, Mächler M, Bolker B, Walker S. Fitting Linear Mixed-Effects Models Using lme4. J Stat Softw 2015; 67: 1–48.

16 Love MI, Huber W, Anders S. Moderated estimation of fold change and dispersion for RNA-seq data with DESeq2. Genome Biol 2014; 15. DOI:10.1186/s13059-014-0550-8.

17 Morton JT, Sanders J, Quinn RA, et al. Balance Trees Reveal Microbial Niche Differentiation. mSystems 2017; 2: e00162–16.

18 McMurdie PJ, Holmes S. phyloseq: An R Package for Reproducible Interactive Analysis and Graphics of Microbiome Census Data. PLOS ONE 2013; 8: e61217.

19 Pianta A, Arvikar S, Strle K, et al. Evidence for Immune Relevance of Prevotella copri, a Gut Microbe, in Patients with Rheumatoid Arthritis. Arthritis Rheumatol Hoboken NJ 2016; published online Nov 18. DOI:10.1002/art.40003.

20 Maeda Y, Kurakawa T, Umemoto E, et al. Dysbiosis Contributes to Arthritis Development via Activation of Autoreactive T Cells in the Intestine. Arthritis Rheumatol 2016; 68: 2646–61.

21 Lo Presti A, Zorzi F, Del Chierico F, et al. Fecal and Mucosal Microbiota Profiling in Irritable Bowel Syndrome and Inflammatory Bowel Disease. Front Microbiol 2019; 10. DOI:10.3389/fmicb.2019.01655.

22 Hofer U. Microbiome: Pro-inflammatory Prevotella? Nat Rev Microbiol 2014; 12: 5–5.

23 Olsen I, Lambris JD, Hajishengallis G. Porphyromonas gingivalis disturbs host–commensal homeostasis by changing complement function. J Oral Microbiol 2017; 9. DOI:10.1080/20002297.2017.1340085.

24 Scher JU, Littman DR, Abramson SB. Review: Microbiome in Inflammatory Arthritis and Human Rheumatic Diseases: MICROBIOME IN RHEUMATIC DISEASES. Arthritis Rheumatol 2016; 68: 35–45.

25 Asquith M, Sternes PR, Costello M-E, et al. HLA Alleles Associated With Risk of Ankylosing Spondylitis and Rheumatoid Arthritis Influence the Gut Microbiome. Arthritis Rheumatol Hoboken NJ 2019; 71: 1642–50.

26 Callahan BJ, McMurdie PJ, Holmes SP. Exact sequence variants should replace operational taxonomic units in marker-gene data analysis. ISME J 2017; 11: 2639–43.

27 Picchianti-Diamanti A, Panebianco C, Salemi S, et al. Analysis of Gut Microbiota in Rheumatoid Arthritis Patients: Disease-Related Dysbiosis and Modifications Induced by Etanercept. Int J Mol Sci 2018; 19. DOI:10.3390/ijms19102938.

28 Chen J, Wright K, Davis JM, et al. An expansion of rare lineage intestinal microbes characterizes rheumatoid arthritis. Genome Med 2016; 8: 43.

29 Mueller RB, Kaegi T, Finckh A, Haile SR, Schulze-Koops H, von Kempis J. Is radiographic progression of late-onset rheumatoid arthritis different from young-onset rheumatoid arthritis? Results from the Swiss prospective observational cohort. Rheumatology 2014; 53: 671–7.

30 Toivanen P, Vartiainen S, Jalava J. Intestinal anaerobic bacteria in early rheumatoid arthritis (RA. Arthritis Res 2002; 4: 5.

